# Sex differences in symptoms following the administration of BNT162b2 mRNA Covid-19 Vaccine in Children below 5 Years of age in Germany (CoVacU5): a retrospective cohort study

**DOI:** 10.1101/2024.03.08.24303999

**Authors:** Jeanne Moor, Nicole Toepfner, Wolfgang C G von Meißner, Reinhard Berner, Matthias B. Moor, Karolina Kublickiene, Christoph Strumann, Cho-Ming Chao

**Affiliations:** CLINTEC Division of Renal Medicine, Karolinska Institutet, Stockholm, Sweden; Department of Pediatrics, Faculty of Medicine and University Hospital Carl Gustav Carus, Technische Universität Dresden, Dresden, Germany; Europäische Fachhochschule, Brühl, Germany; Institute of Family Medicine, University Medical Center Schleswig-Holstein, Campus Luebeck, Luebeck, Germany; LABMED Division of Pathology, Karolinska Institutet, Stockholm, Sweden; Department of Pediatrics, Helios University Medical Center, Witten/Herdecke University, Heusnerstr, Wuppertal, Germany; University Childreńs Hospital, University Medical Center Rostock, University of Rostock, Rostock, Germany; Cardio-Pulmonary Institute (CPI), Universities of Giessen and Marburg Lung Center (UGMLC), Member of the German Center for Lung Research (DZL), Justus Liebig University Giessen, Giessen, Germany

**Keywords:** BNT162b2, children, sex difference, SARS-CoV-2, COVID-19, mRNA vaccine

## Abstract

**Background:** Sex differences exist not only in the efficacy but also in adverse event rates of many vaccines. Here we compared the safety of BNT162b2 vaccine administered off-label in female and male children younger than 5 years in Germany.

**Methods:** This is a retrospective cohort study, in which we performed a post-hoc analysis of a dataset collected through an authentication-based survey of individuals having registered children aged 0-<5 years for vaccination against SARS-CoV-2 in six private practices and/or two lay person-initiated vaccination campaigns. We analyzed the safety profiles of the first 3 doses of 3-10µg BNT162b2. Primary outcome was comparison in frequencies of 4 common post-vaccination symptom categories such as local, general, musculoskeletal symptoms and fever. Data were analyzed according to sex in bivariate analyses and regression models adjusting for age, weight, and dosage. Interaction between sex and BNT162b2 dosage was assessed. An active-comparator analysis was applied to compare post-vaccination symptoms after BNT162b2 versus non-SARS-CoV-2 vaccines.

**Results:** The dataset for the present analysis consisted of 7801 participants including 3842 females (49%) and 3977 males (51%) with an age of 3 years (median, interquartile: 2 years). Among individuals receiving 3µg BNT162b2, no sex differences were noted, but after a first dose of 5 or 10µg BNT162b2, local injection-site symptoms were more prevalent in girls compared to boys. In logistic regression, female sex was associated with higher odds of local symptoms, odds ratio (OR) of 1.33 (95% confidence interval [CI]: 1.15-1.55, p<0.05) and general symptoms with OR 1.21 (95% CI: 1.01-1.44, p<0.05). Following non-BNT162b2 childhood vaccinations, female sex was associated with a lower odds of post-vaccination musculoskeletal symptoms (OR: 0.29, 95% CI: 0.11-0.82, p<0.05). An active comparator analysis between BNT162b2 and non-SARS-CoV-2 vaccinations revealed that female sex positively influenced the association between BNT162b2 vaccine type and musculoskeletal symptoms.

**Conclusions:** Sex differences exist in post-vaccination symptoms after BNT162b2 administration even in young children. These are of importance for the conception of approval studies, for post-vaccination monitoring and for future vaccination strategies.

(German Clinical Trials Register ID: DRKS00028759).

## Introduction

Biological sex introduces a variability in immune responses, and this is therefore pertinent to sex-specific reactions of adaptive immune system following vaccination (1,2). As a consequence, the humoral immune responses of vaccines in humans and in experimental models are more pronounced in females than males as reported for the vaccines against influenza (3,4), hepatitis A or dengue virus (2). The potential underlying mechanisms have been attributed to the immunosuppressive effect of male sex hormones (5,6). In contrast, several observations favor, but still do not universally induce, an increased efficacy in protection from infection in men for some vaccines, including those against SARS-CoV-2 (2,7–9).

Along with the reported sex differences in vaccination efficacy, a plethora of data has revealed sex differences in the rates of adverse events post vaccination (10). For instance, anaphylaxis is 4 times more frequently reported in females than males (8,11). In the case of SARS-CoV-2 vaccines it is opposite, individual undesired sequelae such as myocarditis is more frequently observed in males (12). However, the overall symptoms presentation post-SARS-CoV-2 vaccination remains predominantly reported in females (13–15). In support, vaccine monitoring data and pooled analyses of cross-sectional studies revealed a higher probability of adverse effects in women after received the Pfizer-BioNTech vaccine (Comirnaty®) (16,17). However, further studies are warranted towards SARS-CoV-2 vaccination and implementation of sex-disaggregated statistics (13).

A population that was the last to become eligible for SARS-CoV-2 vaccines were young children. We have previously reported on the overall safety and effectiveness of BNT162b2 in the retrospective CoVacU5 cohort of children vaccinated off-label in Germany (18,19). Such inclusion and assessment represent a unique opportunity to detect otherwise inaccessible dose-response relationships in post-vaccination symptoms.

Even if female sex in adulthood is associated with more frequent adverse events after SARS-CoV-2 vaccination (20), nothing is known in young children. We hypothesized that female sex is associated with greater odds of experiencing adverse symptoms after BNT162b2 vaccination in young children, and that sex specific effects may depend on dose.

To test this hypothesis, we aimed to compare 4 common post-vaccination symptom categories such as local, general, musculoskeletal symptoms and fever after BNT162b2 vaccination in children of both sexes younger than 5 years of age, and we aimed to determine if the safety profile depended on BNT162b2 dose.

## Methods

### Study design

The present study was a post-hoc analysis of the CoVacU5 study. The CoVacU5 study was a retrospective cohort study that assessed symptoms occurring after vaccination with BNT162b2 in children aged less than 5 years old.

### Participants

Participants of the CoVacU5 study were recruited through e-mail addresses of the vaccination registration databases of 21 healthcare institutions and two German vaccination promotion programs “Bildung Aber Sicher” and “U12Schutz”. Inclusion criteria for respondents of the CoVacU5 study survey were having registered a child younger than 5 years old for off-label vaccination with BNT162b2 mRNA vaccine and informed consent of parent or guardian. Exclusion criteria were duplicate entries that had overlaps in all of the four variables age, sex, weight and height, unless the duplicate entry was explained as twins or triplets. Respondents without a correct invitation code were excluded. SARS-CoV-2 vaccinations that were administered before May 2021 or administered using vaccines other than BNT162b2 were also excluded.

### Study procedure

The original survey was available on a web-based REDCap platform (21) from April 15^th^ to May 9^th^ 2022. The survey procedure was pseudonymized and authentication-only as previously described (18). In brief, all eligible potential participants were recruited via the e-mail addresses by which they had previously registered a child aged less than 5 years old for a vaccination with BNT162b2. The invitation e-mails were sent out twice to the eligible individuals, containing an individual access code to allow pseudonymized but authentication-based survey participation only.

The vaccinations with BNT162b2 mRNA vaccine had previously been administered off-label following German law and outside of any study protocol. In addition, the physicians responsible for each vaccinating site were mandated by German law to report each unexpected or severe side effect to the authorities as part of routine medical care.

The study was conducted according to the principles of the Declaration of Helsinki and is reported following the Strengthening the Reporting of Observational Studies in Epidemiology (STROBE) guideline for observational studies. The study protocol was assessed and approved by the Ethics Committee of University of Rostock, Germany (ID: A 2022-0065). The study was registered in the German Clinical Trials Register (ID: DRKS00028759).

### Variables

The survey included close-ended questions gathered demographic information, basic medical data, and timing and dosage of vaccinations with BNT162b2, and all non-SARS-CoV-2 vaccinations in the three months preceding the first BNT162b2 administration. Next, questions covered post-vaccination symptoms occurring after BNT162b2 or non-SARS SARS-CoV-2 vaccinations in 9 symptom categories with close-ended and open-ended follow-up questions to narrow down the characteristics of each individual symptom as described in the initial CoVacU5 study (18).

### Outcomes

The primary outcome of this post-hoc analysis was the sex-specific frequency of symptoms within the four categories of local reaction (injection site), general symptoms (e.g. fatigue, feeling of weakness, general feeling of illness), musculoskeletal symptoms and fever that occurred after first, second and/or third vaccinations with BNT162b2, further stratified by age group and dosage. As secondary analyses, we tested whether an interaction between sex and BNT162b2 dosage was present in association with the primary outcome. Next, we investigated symptoms occurring after non-SARS-CoV-2 vaccinations, and we analyzed symptoms post-BNT162b2 administration in comparison with those occurring after non-SARS-CoV-2 vaccine administration in the period between January 15^th^, 2022 and May 9^th^, 2022.

### Statistical analysis

No power analysis was performed for this study. Analyses were conducted using STATA version 15 and MATLAB R2020a. Categorical variables were compared using Fisher’s exact or Chi-Square test. Continuous variables were assessed using T-test or Wilcoxon rank sum test. Logistic regression models were used with the four symptoms categories as dependent variables, and using sex, age, height, weight and BNT162b2 vaccine dosage as independent variables. Next, sex and dosage were introduced as an interaction term in the models above, and the models with versus without the interaction term were compared using Likelihood Ratio tests. For analyses of non-BNT162b2 vaccines, dosage variable was not included in the models. For the active-comparator analysis comparing post-BNT162b2 symptoms with those occurring after non-BNT162b2 vaccines, we used logistic regression models as above but included vaccine type “BNT162b2” vs. “non-BNT162b2” as a predictor variable. For both bivariate and regression analyses, all p values and 95% confidence intervals underwent adjustment for multiple testing of 4 symptom categories by the Bonferroni method. Statistical significance was assumed at an adjusted p<0.05.

## Results

### Study population

The present analysis included 7801 participants comprising 3842 girls (49%) and 3977 boys (51%) aged 3 years (median, interquartile: 2 years) at the time when receiving their first vaccination with BNT162b2. Demographics, health-related information of participants and the dosage of administered BNT162b2 mRNA vaccines are displayed as sex-disaggregated data in Table 1. The age, height, weight, the chosen BNT162b2 doses, and the fraction of children with comorbidities or long-term medication were similar between the sexes.

**Table 1:** Sample Characteristics, n/N(%)

|  | All<br>n=7801 (100) | Female<br>n=3824 (49) | Male<br>n=3977 (51) |
| --- | --- | --- | --- |
| Age, median (IQR), y | 3 (2-4) | 3 (2-4) | 3 (2-4) |
| Height, median (IQR), cm | 98 (88-105) | 97 (86-104) | 98 (90-105) |
| Weight, median (IQR), kg | 14.5 (12-17) | 14 (12-16) | 15 (12-17) |
| BNT162b2, µg |  |  |  |
| 3 | 2995/14639(20.5) | 1506/7162(21.0) | 1489/7475(19.9) |
| 5 | 6073/14639(41.5) | 2960/7162(41.3) | 3111/7475(41.6) |
| 10 | 5571/14639(38.1) | 2696/7162(37.6) | 2875/7475(38.5) |
| Comorbidities (yes) | 684/7802 (8.8) | 298/3824 (7.8) | 385/3977 (9.7) |
| Long-term medication (yes) | 380/7780 (4.9) | 170/3811 (4.5) | 209/3968 (5.3) |

### Sex-stratified occurrence of post-vaccination symptoms reported after BNT162b2 administration

We assessed the most frequently reported symptom categories of the original CoVacU5 study (18) with sex stratification. The sex-stratified occurrence of local injection site, general, musculoskeletal symptoms and fever after the first administration of BNT162b2 mRNA vaccine are shown in Figure 1. Similarly, symptom categories reported to occur after second or third administration of BNT162b2 mRNA vaccine are shown in Figure 1 and Supplemental Table 1, respectively. The frequency of local injection-site reactions was higher in girls than in boys, and higher at the dose of 10µg BNT162b2. However, there was no significant difference in frequencies between sexes or doses for general, musculoskeletal symptoms and fever after BNT162b2 administration. The underlying numerical data are shown in Supplemental Table 1. Overall, descriptive analysis suggested specific sex differences in post-BNT162b2 symptoms.

**Figure 1.**
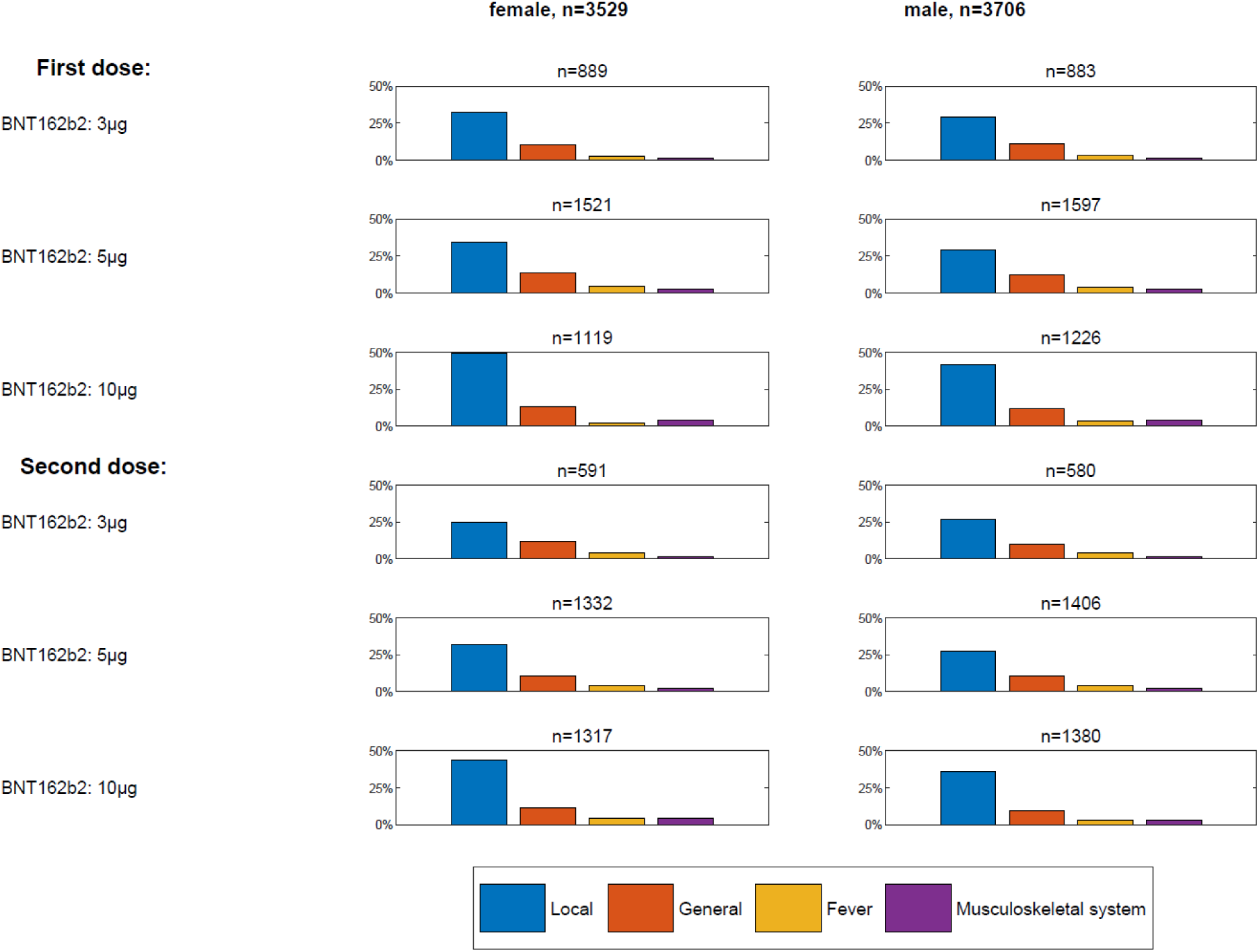
Sex-stratified occurrence of local, general, musculoskeletal symptoms and fever after the first and second administration of BNT162b2 mRNA vaccine.

In logistic regression models adjusted for age, weight and height, female sex was associated with occurrence of local symptoms after BNT162b2 administration (OR: 1.33 [95% CI: 1.15;1.55]). (Table 2) Similarly, female sex was associated with occurrence of general symptoms after BNT162b2 administration (OR: 1.21 [95% CI: 1.01;1.44]). There were no associations between sex and fever or musculoskeletal symptoms (Table 2). The sex-stratified analyses captured associations between BNT162b2 mRNA vaccine dosage and symptoms observed in all symptoms categories (Figure 1, Table 2). However, we found no significant interactions between sex and dosage for symptoms occurring post BNT162b2 vaccination (Table 2).

**Table 2.** Multivariable logistic regression of post-BNT162b2 Symptoms, OR (95% CI)

|  | Local symptoms |  | General symptoms |  | Fever |  | Musculoskeletal symptoms |  |
| --- | --- | --- | --- | --- | --- | --- | --- | --- |
| Model | Model 1 | Model 2 | Model 1 | Model 2 | Model 1 | Model 2 | Model 1 | Model 2 |
| N | 3149 |  | 3142 |  | 3162 |  | 3133 |  |
| Female | 1.33<br>(1.15-1.55) | 1.15<br>(0.78-1.72) | 1.21<br>(1.01-1.44) | 1.06<br>(0.66-1.69) | 0.97<br>(0.73-1.29) | 1.38<br>(0.65-2.94) | 1.23<br>(0.86-1.77) | 0.70<br>(0.16-3.19) |
| 5µg | 1.16<br>(0.92-1.47) | 1.15<br>(0.83-1.60) | 1.39<br>(1.06-1.83) | 1.38<br>(0.94-2.04) | 1.56<br>(1.02-2.39) | 1.95<br>(1.02-3.73) | 2.31<br>(1.03-5.21) | 1.91<br>(0.65-5.61) |
| 10µg | 1.41<br>(1.09-1.82) | 1.19<br>(0.84-1.69) | 1.37<br>(1.00-1.88) | 1.14<br>(0.74-1.75) | 1.18<br>(0.71-1.96) | 1.45<br>(0.71-2.98) | 3.43<br>(1.48-7.95) | 2.35<br>(0.78-7.03) |
| Female X 5µg |  | 1.01<br>(0.65-1.59) |  | 1.01<br>(0.59-1.71) |  | 0.66<br>(0.28-1.54) |  | 1.48<br>(0.30-7.40) |
| Female X 10µg |  | 1.40<br>(0.89-2.22) |  | 1.43<br>(0.82-2.48) |  | 0.68<br>(0.27-1.69) |  | 2.11<br>(0.43-10.29) |
| LR-Test (p-value) | 4.53<br>(0.1038) |  | 3.49<br>(0.1745) |  | 0.97<br>(0.6169) |  | 1.45<br>(0.484) |  |

All models were adjusted for age, weight, height. Models 1 included no interaction terms.
Models 2 included the interaction between sex and dose. Likelihood ratio tests (LR-Tests) determined if inclusion of the interaction term significantly changed the model.

### Sex-specific symptoms occurring after non-BNT162b2 vaccine administration

As a next step, we tested whether a sex effect was present in symptoms occurring after any on-label administration of non-BNT162b2 vaccinations against viruses and bacteria that were performed in the three months prior to a BNT162b2 vaccination, as shown in Supplementary Table 2. In logistic regression models, we found that fever, general or local symptoms were sex-independent (Table 3). However, female sex was negatively associated with musculoskeletal symptoms after non-BNT162b2 vaccine administration (OR: 0.29 [95% CI: 0.11;0.82]). These data indicate that after on-label childhood vaccines, most symptoms categories except musculoskeletal symptoms were not associated with biological sex.

**Table 3.** Multivariable logistic regression of post-Non-BNT162b2 Symptoms, OR (95% CI)

|  | Local<br>symptoms | General<br>symptoms | Fever | Musculoskeletal<br>symptoms |
| --- | --- | --- | --- | --- |
| Observations | 1473 | 1473 | 1473 | 1473 |
| n/N(%) | 380 (25.5) | 388 (26) | 256 (17.2) | 21 (1.4) |
| Female | 1.16 (0.91-1.48) | 1.00 (0.78-1.28) | 0.85 (0.64-1.14) | 0.29 (0.11-0.82) |

All models were adjusted for age, weight, height.

### Active comparator analysis of symptoms occurring after BNT162b2 mRNA vaccine versus other vaccine administration in the same population

Further, we aimed to compare the symptoms occurring in a cohort of 4570 children who had received at least one BNT162b2 mRNA vaccine within the last 3 months with those occurring in 1473 individuals of the same population after receiving the non-BNT162b2 vaccinations listed in Supplementary Table 2. Using logistic regression analysis of all vaccinations combined and adjusting for age, weight and height, we found that BNT162b2 mRNA vaccine was associated with higher odds of symptoms of the local injection-site and the musculoskeletal category, but at lower odds of general symptoms or fever (Table 4). In this analysis, female sex was associated with increased odds of post-vaccination local symptoms (OR: 1.35 [95% CI: 1.17-1.56], p<0.05) but not with any of the other three symptoms categories (Table 4). Secondary analyses revealed an interaction between vaccine type and sex for the musculoskeletal category only (interaction BNT162b2 mRNA vaccine and female sex: OR 4.01 [95% CI: 1.01;16.02], Likelihood ratio test p=0.021), (current Table 5; final Table 4). Overall, these results indicate that few post-vaccination symptoms showed an association with sex in the combined dataset of BNT162b2 and non-BNT162b2 vaccination, with the exception that after non-BNT162b2 vaccinations the probability of musculoskeletal symptoms was negatively affected by female sex.

**Table 4.** Comparison of symptoms occurring after BNT162b2 and after non-BNT162b2 vaccinations.

|  | Local symptoms |  | General symptoms |  | Fever |  | Musculoskeletal symptoms |  |
| --- | --- | --- | --- | --- | --- | --- | --- | --- |
| Observations | 5918 |  | 5904 |  | 5962 |  | 5894 |  |
| *Frequency distributions* |  |  |  |  |  |  |  |  |
| BNT162b2<br>n/N(%) | 1808/4520 (40.0) |  | 874/4506 (19.4) |  | 305/4570 (6.7) |  | 193/4496 (4.3) |  |
| non-BNT162b2:<br>n/N(%) | 380/1490 (25.5) |  | 388/1490 (26.0) |  | 256/1490 (17.2) |  | 21/1490 (1.4) |  |
| *Multivariable logistic regression* |  |  |  |  |  |  |  |  |
| BNT162b2<br>vs non-BNT162b2 | 1.68<br>(1.41-2.01) | 1.58<br>(1.23-2.03) | 0.77<br>(0.64-0.93) | 0.73<br>(0.57-0.95) | 0.42<br>(0.33-0.54) | 0.40<br>(0.29-0.56) | 2.55<br>(1.39-4.67) | 1.49<br>(0.72-3.05) |
| Female sex | 1.35<br>(1.17-1.56) | 1.22<br>(0.90-1.67) | 1.12<br>(0.94-1.32) | 1.04<br>(0.76-1.42) | 0.96<br>(0.76-1.22) | 0.90<br>(0.62-1.29) | 1.19<br>(0.82-1.71) | 0.34<br>(0.09-1.27) |
| Female sex<br>X<br>BNT162b2 |  | 1.13<br>(0.79-1.60) |  | 1.10<br>(0.77-1.59) |  | 1.13<br>(0.70-1.81) |  | 4.01<br>(1.01-16.02) |
| LR-Test<br>(p-value) | 0.79<br>(>0.999) |  | 0.48<br>(>0.999) |  | 0.41<br>(>0.999) |  | 7.80<br>(0.021) |  |

All models were adjusted for age, weight, height. Models 1 included no interaction terms. Models 2 included the interaction between sex and dose. Likelihood ratio tests (LR-Tests) determined if inclusion of the interaction term significantly changed the model.

## Discussion

In the present study, we investigated the sex differences in post-vaccination symptoms in young children using a large retrospective cohort including COVID-19 and non-COVID-19 vaccinations in Germany. Our study supports the notion that biological sex plays an important role in the presentation of post-vaccination symptoms. The study findings demonstrate that post-vaccination symptoms predominantly occur in female children even at very young age far before puberty. Our main findings were that i) female sex was significantly associated with occurrence of local symptoms and general symptoms after BNT162b2 administration. ii) Female sex was associated with increased odds of post-vaccination local symptoms among all vaccinations, regardless of type. iii) Female sex was associated with lower odds of musculoskeletal symptoms occurring after non-BNT162b2 childhood vaccine administration.

The increased probability of local injection-site symptoms in female compared to male sex after any type of vaccination in this pediatric population is in accordance with previous reports on various vaccines (22,23). The current BNT162b2 assessment in young children is important as previous reports on safety of BNT162b2 administration in this age group either did not stratify for sex (18,24,25) or found no sex differences (26).

The mechanism underlying the increased odds of local and general symptoms in young girls receiving BNT162b2 is not clear. For vaccination in general, such a sex difference is suspected to be multifactorial and related to pain sensitivity, hypersensitivity reactions or most likely hormonal factors (22). The observed sex difference for BNT162b2 may also be in part attributable to the innate immune system, as females are known to have stronger innate immune responses than males, sometimes with non-specific adverse vaccine effects (27–29). The sex difference observed in the present study are less likely to be caused by inherent differences in the adaptive immune responses, as BNT162b2-induced SARS-CoV-2 neutralizing antibody titers were comparable between the sexes in this age group according to BNT162b2 manufacturer data (26). Additionally, the effects were less likely to depend on hormonal factors as the assessed age group was far prior puberty. In young children, the hormone environment may have only limited influence on the immune system compared to what could be expected in adults of reproductive age, and hence the obtained findings could be related to the intrinsic hormone-independent sex-specific effects.

The strengths of this study include the big sample size of the CoVacU5 dataset and the focus on a very young age group. Sex-stratified analyses in this group is often neglected despite the potential insight to differentiate hormone-dependent from actual sex-specific effects.

The present study also has limitations that need to be acknowledged. These include that no causal effects are investigated between vaccination and symptoms due to the observational design of the study, and that the symptom reports by legal guardians are of subjective nature and underlie both recall bias and potentially depend on the gender of the reporting person which was not reported. Finally, inflammatory post-vaccination symptoms including but not limited to myocarditis that are more frequent in male than female adolescents (30) were not assessed in this study due to the low number of events, and they are of importance and warrant additional investigation in young children.

In conclusion, the finding of increased odds of local and general symptoms in young girls compared to boys is apparent in young children after administration of COVID-19 vaccine.

This observation has implications for present and future conceptions of approval studies as well as post-vaccination monitoring and the design of future mRNA applications. The choice of the pro-inflammatory mRNA vaccine delivery vehicle (i.e., types of lipids used for encapsulation) may thus be reevaluated in the future due to sex differences in post-vaccination symptoms affecting even very young children far before puberty.

## Author contributions

Dr Strumann had full access to all of the data in the study and takes responsibility for the integrity of the data and the accuracy of the data analysis. Drs Strumann and Chao are co– senior authors.

Concept and design: J. Moor, Strumann, M.B. Moor., Chao

Acquisition, analysis, or interpretation of data: J. Moor, Töpfner, von Meißner, Berner, M.B. Moor., Kublickiene, Strumann, Chao

Drafting of the manuscript: J. Moor, M.B. Moor., Kublickiene

Critical revision of the manuscript for important intellectual content: J. Moor, Töpfner, von Meißner, Berner, M.B. Moor., Kublickiene, Strumann, Chao

Statistical analysis: Strumann

Administrative, technical, or material support: Töpfner, Berner, Chao. Supervision: Kublickiene, Strumann, Chao

## Acknowledgements

The authors want to thank all participating vaccination institutions, legal guardians, and children. We are thankful to the initiatives U12Schutz and Bildung Aber Sicher for help with recruitment of respondents.

## Data availability

A restricted dataset is available from the corresponding author on request. Access to the full dataset can be requested upon approval by the Ethics Committee of University of Rostock, as it contains potentially identifying patient information.

## Registration

The present work was registered at the German Clinical Trials Register ID: DRKS00028759.

---

**Supplemental Table 1.** Post-vaccination symptoms after BNT162b2 vaccine, n/N (%)

| Dose | Symptoms | First Vaccination |  |  | Second Vaccination |  |  | Third Vaccination |  |  |
| --- | --- | --- | --- | --- | --- | --- | --- | --- | --- | --- |
|  |  | Female | Male | p-val<sup>a</sup> | Female | Male | p-val<sup>a</sup> | Female | Male | p-val<sup>a</sup> |
| 3µg | Local | 286/883 (32.4) | 252/874 (28.8) | 0.4233 | 145/588 (24.7) | 153/574 (26.7) | >0.999 | 3/26 (11.5) | 9/26 (34.6) | 0.1931 |
|  | General | 92/881 (10.4) | 94/868 (10.8) | >0.999 | 69/587 (11.8) | 55/569 (9.7) | >0.999 | 1/26 (3.8) | 2/26 (7.7) | >0.999 |
|  | Fever | 23/881 (2.6) | 27/868 (3.1) | >0.999 | 24/587 (4.1) | 22/569 (3.9) | >0.999 | 0/26 (0) | 0/26 (0) | - |
|  | Musculosk | 11/877 (1.3) | 13/868 (1.5) | >0.999 | 8/584 (1.4) | 8/571 (1.4) | >0.999 | 0/25 (0) | 0/26 (0) | - |
| 5µg | Local | 522/1511 (34.5) | 461/1590 (29) | **0.0036** | 423/1324 (31.9) | 388/1401 (27.7) | 0.0608 | 38/107 (35.5) | 39/108 (36.1) | >0.999 |
|  | General | 203/1508 (13.5) | 194/1589 (12.2) | >0.999 | 142/1322 (10.7) | 151/1400 (10.8) | >0.999 | 10/107 (9.3) | 14/108 (13) | >0.999 |
|  | Fever | 65/1508 (4.3) | 60/1589 (3.8) | >0.999 | 56/1322 (4.2) | 60/1400 (4.3) | >0.999 | 8/107 (7.5) | 4/108 (3.7) | 0.9129 |
|  | Musculosk | 39/1505 (2.6) | 39/1586 (2.5) | >0.999 | 28/1319 (2.1) | 35/1396 (2.5) | >0.999 | 5/107 (4.7) | 1/108 (0.9) | 0.3814 |
| 10µg | Local | 548/1111 (49.3) | 504/1217 (41.4) | **0.0005** | 569/1308 (43.5) | 493/1374 (35.9) | **0.0002** | 92/258 (35.7) | 112/267 (41.9) | 0.5578 |
|  | General | 142/1106 (12.8) | 143/1214 (11.8) | >0.999 | 150/1304 (11.5) | 132/1372 (9.6) | 0.4519 | 37/258 (14.3) | 31/267 (11.6) | >0.999 |
|  | Fever | 23/1106 (2.1) | 44/1214 (3.6) | 0.1059 | 54/1304 (4.1) | 43/1372 (3.1) | 0.6544 | 16/258 (6.2) | 13/267 (4.9) | >0.999 |
|  | Musculosk | 44/1105 (4) | 47/1208 (3.9) | >0.999 | 59/1302 (4.5) | 38/1366 (2.8) | 0.0632 | 7/258 (2.7) | 15/264 (5.7) | 0.3658 |

<sup>a</sup>Adjusted for multiple testing by Bonferroni correction.

**Supplemental Table 2.** Non-BNT162b2 vaccines administered since January 15^th^ 2022, n (%)

|  | All<br>n=4570<br>(100) | Female<br>n=2251<br>(49.3) | Male<br>n=2319<br>(50.7) | p-val<sup>a</sup> |
| --- | --- | --- | --- | --- |
| Non-BNT162b2 vaccines (yes) | 1490 (32.6) | 718 (31.9) | 772 (33.3) | >0.999 |
| Influenza | 540 (11.8) | 254 (11.3) | 286 (12.3) | >0.999 |
| Meningococcal | 476 (10.4) | 245 (10.9) | 231 (10) | >0.999 |
| Measles/mumps/rubella<br>with/without chickenpox | 387 (8.5) | 195 (8.7) | 192 (8.3) | >0.999 |
| Tetanus/diphtheria/pertussis and/or<br>pediatric polio | 364 (8) | 181 (8) | 183 (7.9) | 0.279 |
| Hepatitis A/B | 159 (3.5) | 71 (3.2) | 88 (3.8) | >0.999 |
| Human papillomavirus | 4 (0.1) | 1 (0) | 3 (0.1) | >0.999 |
| Other | 266 (5.8) | 127 (5.6) | 139 (6) | >0.999 |

<sup>a</sup>Adjusted for multiple testing by Bonferroni correction.

